# Acute Demyelinating Encephalomyelitis (ADEM) in COVID-19 infection: A Case Series

**DOI:** 10.1101/2020.07.15.20126730

**Authors:** Micheala McCuddy, Praful Kelkar, Yu Zhao, David Wicklund

## Abstract

**Objective:** To report three patients infected with COVID-19 with severe respiratory syndrome requiring intubation, who developed acute demyelinating encephalomyelitis (ADEM).

**Method:** Patient data were obtained from medical records from the North Memorial Health Hospital, Robbinsdale, MN, USA

**Results:** Three patients (two men and one woman, aged 38 - 63) presented with fatigue, cough and fever leading to development of acute respiratory distress syndrome secondary to COVID-19 infection requiring intubation and ventilatory support. Two patients were unresponsive, one with strong eye deviation to the left and the third patient had severe diffuse weakness. MRI in all patients showed findings consistent with ADEM. CSF showed elevated protein in all patients with normal cell count and no evidence of infection, including negative COVID-19 PCR. All three of the patients received Convalescent plasma therapy for COVID-19. All patients were treated with intravenous corticosteroids and improved, although two responded minimally. Two patients treated with IVIG showed no further improvement.

**Conclusion:** Neurological complications from COVID-19 are being rapidly recognized. Our three cases highlight the occurrence of ADEM as a postinfectious/immune mediated complication of COVID-19 infection, which may be responsive to corticosteroid treatment. Early recognition of this complication and treatment is important to avoid long term complications.

## Introduction

Acute Disseminated Encephalomyelitis (ADEM) is a rare, immune-mediated, demyelinating disorder of the central nervous system. It is usually monophasic, characterized by acute encephalopathy with neurologic deficits and brain MRI findings consistent with multifocal demyelination. ADEM is a postinfectious/immune mediated complication usually following infections, with the highest incidence in the pediatric population. Dating back to the 18th century, it has been associated primarily with viral pathogens including measles, mumps and rubella. ^1^ More recently, it has been described in association with Middle East respiratory syndrome coronavirus (MERS) and Coronavirus OC43.^2^

With the backdrop of t0he current COVID-19 pandemic, various neurologic complications are being reported. ^3^ ADEM secondary to COVID-19 has been described in only one prior patient to date. ^4^ Similar cases of brain and spine demyelinating were also reported. ^5,6^ We present a series of 3 adult patients from one medical center with severe COVID-19 infection with acute respiratory failure requiring intubation, who developed neurological complications with MRI changes indicative of ADEM.

## Method

Patient data were obtained from medical records from the North Memorial Hospital, Robbinsdale, MN, USA.

## Results

We report three adult patients who developed ADEM as a complication of severe COVID-19 infection (See Table 1). Two were males and one female with ages between 38-70 years. The common concomitant illness was diabetes mellitus. All 3 patients presented with fatigue, cough and fever leading to development of acute respiratory distress syndrome secondary to COVID-19 infection requiring ventilatory support. They were intubated for 16-32 days and were diagnosed with ADEM during the third week of the illness. Neurological presentation consisted of encephalopathy post-extubation with unresponsiveness in two patients, with eye deviation to the left in one. The third patient developed severe diffuse weakness with hyperreflexia.

**Table 1.** Clinical characteristics, MRI and laboratory findings and outcome in three patients with Covid-19 infection related ADEM.

|  | **Patient 1** | **Patient 2** | **Patient 3** |
| --- | --- | --- | --- |
| **Age/Gender** | About 40, F | About 60, M | About 70, F |
| **Concomitant Illness** | DMII, HTN, obesity and pregnancy at 30wks gestation. | DMII, HTN, CKD3 asthma. | DMII, HTN,HLD, CKD2, morbid obesity, peripheral neuropathy and glaucoma |
| **Presentation and Initial Course** | Presented with cough, chest pain, fever up to 101°F and worsening shortness of breath.<br>Admitted for acute respiratory failure, ultimately requiring emergent c-section and intubation.<br>Diffuse weakness noted post-extubation. | Admitted for poor appetite, fever and acute respiratory failure<br>Neurology consulted for unresponsiveness off of sedation and difficulty weaning, with strong gaze deviation to the left. | Presented with decreased appetite, fatigue, generalized weakness and lethargy and cough. Developed hyperglycemia, AKI, atrial fibrillation, sepsis and respiratory failure leading to intubation.<br>Neurology consulted for unresponsiveness off sedation. |
| **Days intubated for COVID-19** | 16 | 32 - tracheostomy, remains ventilator dependent | 26 - remains on ventilator support, weaning. |
| **Diagnosis of ADEM on day # of hospitalization** | 22 | 20 | 16 |
| **Significant exam findings at time of ADEM diagnosis.** | Patient plegic in legs bilaterally. Significant, symmetric weakness in the upper extremity. DTRs brisk. No clonus or Babinski. Mental status and sensation intact. | Unresponsive. Eyes with leftward deviation Corneal and gag reflex intact. No spontaneous limb movements. Reflexes reduced.. | Unresponsive to verbal stimuli.. Withdraws to pain slightly. Upgoing toes. |
| **MRI findings** | **MRI Brain (with/without contrast):**<br>Multiple T2 hyperintense lesions with restricted diffusion involving the corpus callosum, bilateral cerebral white matter, right pons and in the bilateral ventral medulla. Some enhancement associated with the largest lesion, which is a 3 cm lesion in the body of the corpus callosum. No hemorrhage.<br>**MRI of cervical/thoracic/lumbar spine** unremarkable. | **MRI Brain (no contrast due to renal failure):**<br>Several T2 hyperintense lesions, many with restricted diffusion, in cerebral white matter as well as deep bilateral cerebellum. No hemorrhage.<br>**Follow-up MRI 5 days later:** No significant change, slightly less restricted diffusion. | **MRI Brain (with/without contrast):** Several T2 hyperintense lesions, most with restricted diffusion, in deep cerebral white matter and corpus callosum as well as left brachium pontis. Basal ganglia and thalamus spared. Minimum enhancement and no hemorrhage.<br>**Follow up MRI brain with and without contrast 8 days later:** The intensity of restricted diffusion and T2 signal is similar though slightly improved, although the size of the foci is stable, with the exception of a small focus of restricted diffusion within the deep left cerebellar white matter which has improved. |
| **CSF Analysis** | Protein – 95mg/dL<br>Glucose – 85mg/dL<br>Cell Count - WBC: 2/uL<br>Oligoclonal Bands- 0<br>Meningitis/encephalitis panel, lyme, MS panel, VDRL all negative.<br>*\*Experimental COVID CSF PCR negative.* | Protein – 55mg/dL<br>Glucose – 112mg/dL<br>Cell Count - WBC:1/uL<br>Oligoclonal Bands - 0<br>CSF Culture and MS panel, Lyme and VDRL negative.<br>*\*Experimental COVID CSF PCR negative.* | Protein – 63mg/dL<br>Glucose – 87mg/dL<br>Cell Count - WBC:0/uL<br>Oligoclonal Bands - 3<br>Meningitis/encephalitis panel, CSF culture, MS panel, Lyme and VDRL negative<br>*\*Experimental COVID CSF PCR negative.* |
| **Treatment** | Decadron 20mg IV x 5days and 10mg x 5days<br>Also received Hydroxychloroquine, Zinc and Convalescent Plasma Therapy | IV Solumedrol 1 g/d x 5 days and IVIG 25g/day x 3 days<br>Received Convalescent plasma therapy | IV Solumedrol 1g/d x 5 days and IVIG 25g/day x 3 days<br>Received Convalescent plasma therapy |
| **Outcome** | **50 days after admission:**<br>Improving. Partial return of strength in upper extremities core. Regaining some function in distal lower extremities. Cognition intact. | **35 days after admission:**<br>Interval improvement on imaging. Clinically, not opening eyes, unresponsive to verbal/painful stimuli and no purposeful movements. Remains on ventilator with tracheostomy. | **26 days after admission:**<br>Interval improvement on imaging. Spontaneously opens eyes, blinks to confrontation, flexion withdrawal vs. decorticate posturing in upper extremity. Withdraws lower extremity to stimuli. Weaning from ventilator. |

Brain MRI (figure 1) in all three cases showed multiple T2 hyperintense lesions distributed predominantly in the white matter, many showing restricted diffusion. These were bilateral and asymmetric; small, round to oval, and with somewhat indistinct margins. The lesions were in the deep and periventricular white matter with involvement of corpus callosum and brainstem. Minimal enhancement was seen in a few lesions. There was little if any disease in the basal ganglia or thalami. There was no associated hemorrhage.

**Figure 1.**
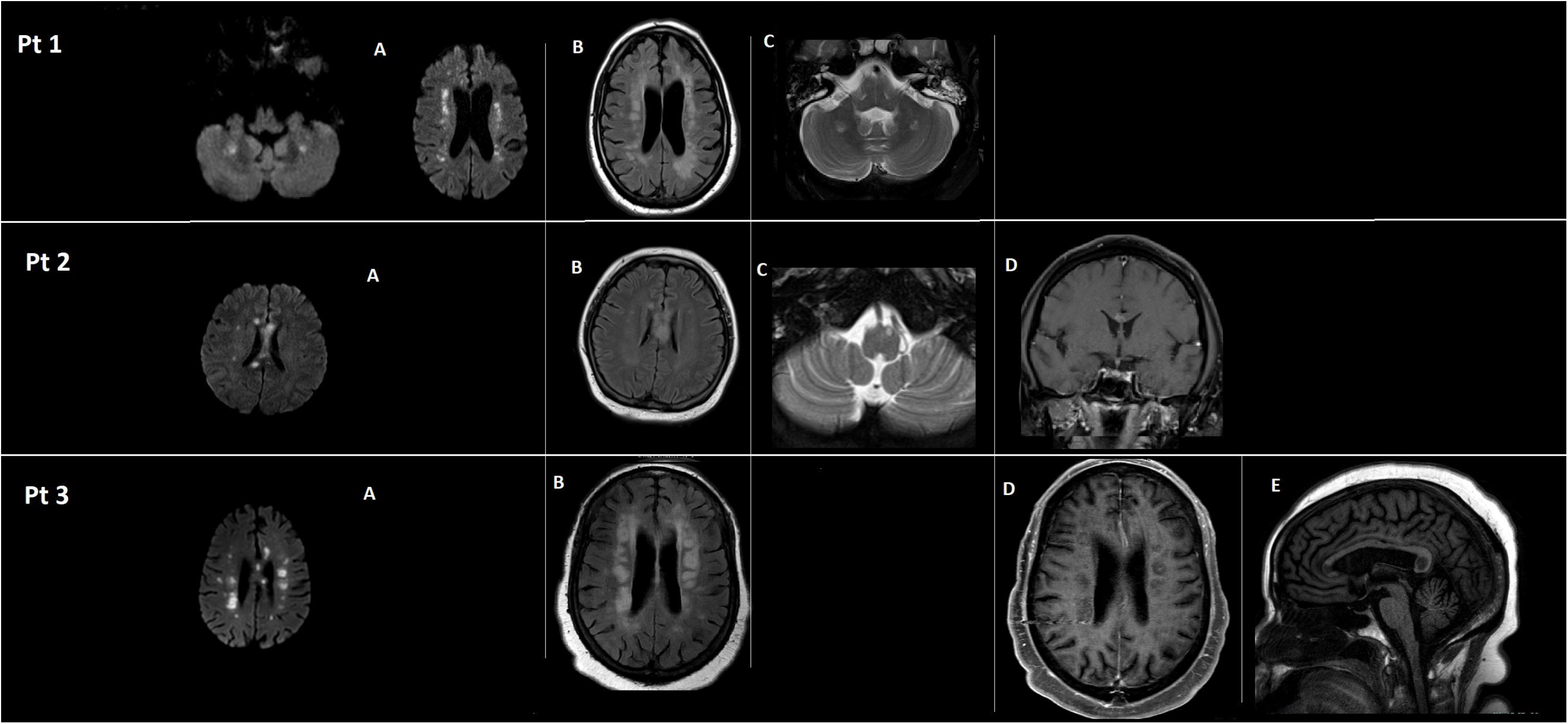
MRI findings in three patients showing multifocal T2 hyperintense lesions with restricted diffusion involving the corpus callosum, bilateral cerebral white matter, brainstem with some enhancement. A = Diffusion (DWI); B = T2 FLAIR; C = Fat-suppressed T2; D = Post-contrast T1; E = Non-contrast T1.

CSF showed elevated protein 55-95 mg/dL with normal cell count. Cultures and encephalitis/meningitis panels, oligoclonal bands and IgG index were negative in all cases. CSF COVID-19 PCR was run on an experimental basis at the Mayo Clinic Laboratory and was negative in all cases, whereas serum PCR was positive for COVID-19 in all.

All three patients received convalescent plasma treatment for COVID-19, and IV steroids (solumedrol in 2 and dexamethasone in 1) and two received IVIG. Following steroid infusions Patient 1 improved, patients 2 and 3 both demonstrated mild improvement on MRI findings, although their neurologic change was minimal. Both also received subsequent IVIG dosed at 0.4g/kg for 3 days with no further improvement.

## Discussion

There are increasing reports of neurologic manifestations of COVID-19 infection with the incidence of CNS symptoms shown to be as high as 40% in patients with severe infection.^3, 7^ Ischemic strokes, encephalopathy, meningitis, encephalitis, Guillain Barre syndrome or Miller-Fisher syndrome and acute hemorrhagic necrotizing encephalopathy have been described.^3, 7, 8^

ADEM due to Covid-19 was previously described in one patient ^4^ and similar MRI findings suggestive of demyelination were reported in two recent cases. ^5, 6^ We now describe a series of three patients with severe Covid-19 infection who developed ADEM. All these patients were intubated for respiratory failure. Two patients were unresponsive and the third patient had severe diffuse weakness.

Magnetic resonance is the preferred imaging modality in assessment of suspected cases. All of the patients had unremarkable CT scans. MRI in all of these patients showed findings indicative of ADEM. Characteristic T2 hyperintense lesions were small, round to oval with somewhat indistinct margins, and many had restricted diffusion. The distribution and morphology of the lesions were not typical for acute embolic infarcts. None had history of preexisting demyelinating disorder such as multiple sclerosis, which can have a similar appearance. The relative lack of enhancement and negative CSF panels argue against common infectious encephalitis, with or without abscess. Atypical features in this series might include involvement of the corpus callosum and relative lack of signal abnormalities in deep gray nuclei.

All three patients underwent thorough investigation to rule out other causes of CNS involvement. CSF in all 3 patients showed elevated protein with normal WBC count and negative oligoclonal bands.They all had negative CSF cultures, negative meningitis/encephalitis panels, negative MS panels and CSF negative for lyme and syphilis. All patients also had negative COVID-19 PCR of CSF (done on an experimental basis at Mayo Clinic) and thus there was no evidence of direct CNS involvement by the SARS-CoV-2 coronavirus. Findings were overall consistent with an immune-mediated process.

All three patients received IV steroids and two also received IVIG for ADEM. Patient 1 had significant improvement with return of strength in upper extremities and improved function in lower extremities. Patients 2 and 3 both demonstrated mild interval improvement on MRI findings after IV Solumedrol, but minimal neurologic changes, and no further changes after IVIG treatment. A common comorbidity in all three patients was diabetes mellitus.

The acute encephalopathy and multifocal neurologic deficits seen in ADEM have the potential to compound the morbidity in patients with severe COVID-19 infections, prolong hospitalizations and lead to long-term neurologic deficits. It is thought that the pro-inflammatory state induced by the cytokine storm may be responsible for the glial cell activation and subsequent demyelination. As such, the accurate diagnosis and timely treatment of ADEM in these patients is of significant clinical importance. There are no randomized studies for ADEM treatment. When treated with high dose corticosteroids (Methylprednisolone is considered first-line) in the acute phase, ADEM has a reported 50-80% rate of full recovery. IVIG and plasma exchange are considered second line therapies.^1^ Use of plasma exchange is limited by the potential to exacerbate hemodynamic instability, risk of infection and potential to decrease the protective antibodies that may help to fight COVID-19. IVIG can increase the risk of thromboembolic adverse events in COVID-19 patients, who already are at increased risk for hypercoagulability.^9^.

The question of the safety of corticosteroid use in COVID-19 remains a matter of debate. There is concern that steroid use can augment viral replication, while steroids also have the capacity to modulate the inflammatory response responsible for many of the severe complications of this infection. Given lack of current clinical evidence for or against steroid use, best clinical judgement should be used when treating COVID-19 associated ADEM and corticosteroid therapy should not be disregarded when profound neurologic deficits have the potential to undermine all other treatment efforts. None of the patients in our series had any adverse effects from IV steroid treatment.

In summary, clinicians need to be aware of the potential for ADEM in acutely ill COVID-19 patients. Unexplained unresponsiveness, weakness or development of focal central neurological signs should alert clinicians to this possibility. MRI findings are characteristic and helpful in making the diagnosis. Treatment with corticosteroids should be considered in these patients.

## Data Availability

All the data is maintained by the authors.

